# Microbiota of Chronic Prostatitis/Chronic Pelvic Pain Syndrome are Distinct from Interstitial Cystitis/Bladder Pain Syndrome

**DOI:** 10.1101/2021.03.04.21252926

**Authors:** Bryan White, Michael Welge, Loretta Auvil, Matthew Berry, Colleen Bushell, Anthony J. Schaeffer, Ryan E. Yaggie, James W. Griffith, David J. Klumpp

## Abstract

Urologic chronic pelvic pain syndrome patients include men chronic prostatitis/chronic pelvic pain syndrome (CP/CPPS) and patients, mainly women, with interstitial cystitis/bladder pain syndrome (IC/BPS or IC). CP/CPPS is marked by severe chronic pelvic pain of unknown etiology that is differentially associated with prostatic inflammation. Microbes are known to modulate sensory responses, and microbiota are increasingly understood to drive normal biological processes and pathogenesis, including inflammation. Recent studies have linked fecal dysbiosis with chronic pelvic pain in IC/BPS, suggesting a role for microbiota in modulating UCPPS pain. Similarly, dysbiosis has been reported in CP/CPPS patients, but the relationship between with the dysbiosis of IC/BPS patients is unclear. Here, we characterized the fecal microbiota of men with CP/CPPS and women and men with IC/BPS. Similar to recent reports, we identified fecal dysbiosis in men with CP/CPPS relative to healthy controls among specific phyla and overall differences in diversity and richness. Interestingly, we also observed differences between CP/CPPS microbiota and IC/BPS microbiota that were not likely due to sex differences. These findings suggest that CP/CPPS is marked by changes in the gut microbiome, but these changes differ from IC/BPS. Taken together, UCPPS appears associated with distinct dybioses among CP/CPPS and IC/BPS, raising the possibility of distinct contributions to underlying pelvic pain mechanisms and/or etiologies.

## INTRODUCTION

Urologic chronic pelvic pain syndromes (UCPPS) cause significant morbidity for millions in the U.S., but underlying mechanisms are unclear, etiologies are unknown, and effective therapies are lacking [1]. Chronic prostatitis/chronic pelvic pain syndrome (CP/CPPS or “prostatitis”) is characterized by severe pelvic pain and is associated with co-morbid anxiety and depression [2, 3]. Similarly, interstitial cystitis/bladder pain syndrome (IC/BPS or “IC”) is a debilitating condition of severe pelvic pain that afflicts women disproportionately and is often accompanied by voiding dysfunction and co-morbid anxiety and depression [4-6]. Because UCPPS mechanisms and therapies remain elusive, NIDDK launched their flagship effort in pelvic pain, the Multi-Disciplinary Approach to the Study of Chronic Pelvic Pain Research Network (MAPP, [7, 8]) supporting deep phenotyping of UCPPS patients and mechanistic studies in clinically relevant models. Historically, UCPPS were viewed as end-organ conditions of the prostate or bladder, but MAPP findings suggest a centralized component of pelvic pain from fMRI signatures of UCPPS, altered cognitive function, and patient subsets exhibiting differential pain “widespreadness” [9]. Together, these findings suggest a systemic influence on UCPPS.

The microbiome shapes normal biology and pathogenesis [10-12], and this is increasingly understood to extend to urology. Pioneering work by Brubaker and Wolfe identified a normal human urinary microbiome that may shift and thereby contribute to urologic disease, and MAPP is currently evaluating microbiota UCPPS [13, 14]. However, given that bowel-bladder organ crosstalk influences the bladder via visceral convergence [15-17], we hypothesized that gut microbes influence pelvic pain. Indeed, we observed fecal dysbiosis in women with IC [18], supporting the hypothesis that gut microbiota influence UCPPS. Using 16S sequencing of fecal stool from controls and IC patients, we identified many significant differences at the phylum level. In silico metagenome analyses correlated with stool metabolomics in implicating altered lipid metabolism. Enhanced random forest identified OTUs that co-varied with patient symptom scores; these were validated and identified species that were deficient in IC pelvic pain (DIPP).

In CP/CPPS, Shoskes and colleagues defined microbiota in rectal stool [19]. They identified altered microbiota among CP/CPPS stool relative to controls. CP/CPPS microbiota were characterized by reduced alpha diversity compared to healthy stool, and several taxa exhibited altered abundance. These differences included numerous taxa that were of decreased abundance, reminiscent of DIPP species in IC fecal microbiota [18, 19]. Together, these findings in IC/BPS and CP/CPPS indicate gut dybiosis in UCPPS that may contribute to or result from the pathogenesis of chronic pelvic pain.

Here, we performed 16S analyses on fecal stool of CP/CPPS patients, relative to healthy controls and compare these findings with fecal microbiota of IC/BPS. Consistent with prior studies, we observed altered microbiota among CP/CPPS patients as well as differences with IC/BPS microbiota. These results confirm dysbiosis in CP/CPPS patients and suggest that dysbiosis differs between CP/CPPS and IC/BPS that may reflect differences in pathogenesis among distinct UCPPS.

## MATERIALS AND METHODS

### Participants

Participants included healthy controls, UCPPS (CP/CPPS and IC/BPS), and clinical control groups of patients with over-active bladder (OAB), lower urinary tract symptoms (LUTS), and major-depressive disorder (MDD). Participants were recruited from Urology Clinic at Northwestern Memorial Hospital, the Department of Medical Social Sciences (MDD patients), CT.gov, and posted bulletins. Some participants identified in the Urology Clinic were also participants in the NIDDK MAPP study; these participants were recruited following and independent of their consent to participate in MAPP. Prospective participants were screened a trained Research Coordinator in the clinic and provided written, informed consent by signature of a consent form approved by the Northwestern University Institutional Review Board (NU IRB protocol STU00055668). Alternatively, prospective participants identified initially through CT.gov or responses to posted bulletins were similarly screened by phone, including confirming diagnosis by a licensed clinician, and then provided oral consent; written consent forms were delivered to these participants with questionnaires and stool collection materials.

### Inclusion/exclusion criteria

For urologic patients, eligible patients were 18 years old or older, reporting unpleasant sensation of pain, pressure or discomfort, perceived to be related to the bladder and/or pelvic region or clinically diagnosed with any of the following: IC, Painful Bladder Syndrome, Bladder Pain Syndrome, CP/CPPS, LUTS or OAB. Patients were excluded if they met the following criteria: on-going symptomatic urethral stricture, on-going neurological disease or disorder affecting the bladder or bowel fistula, history of cystitis caused by tuberculosis, radiation therapy or Cytoxan/cyclophosphamide therapy, had augmentation cystoplasty or cystectomy, had an active autoimmune or infectious disorder (such as Crohn’s Disease or Ulcerative Colitis, Lupus, Rheumatoid Arthritis, Multiple Sclerosis, or HIV), had a history of cancer (with the exception of skin cancer), had any psychiatric or medical comorbidities that would interfere with study participation (e.g. dementia, psychosis, upcoming major surgery, lupus, active heart failure, diabetes), had a UTI and/or a positive urine culture in the prior 6 weeks and/or was taking antibiotics or had done so in prior last 3 months. For MDD patients, eligible patients were 18 years old or older, having a current diagnosis of MDD. Patients were excluded if they met the following criteria: currently in remission or has recovered from major depressive disorder, has a substance use disorder in the past 6 months, had been diagnosed with any bipolar disorder, had been diagnosed with any psychotic disorder, had been diagnosed with any severe cognitive impairment or dementia, had a history of cancer (with the exception of skin cancer), had any psychiatric or medical comorbidities that would interfere with study participation (e.g. dementia, psychosis, pending major surgery, lupus, active heart failure, diabetes, etc), had a UTI and/or a positive urine culture in the prior 6 weeks and/or was taking antibiotics or has done so in the prior 3 months.

### Sample handling and 16S analyses

Samples were received and processed as previously described [18]. Briefly, patients were provided with a current consent approved by the Northwestern Institutional Review Board, a home stool collection kit per HMP protocols, a demographic and clinical questionnaire, and a male-specific or female-specific Genitourinary Pain Index questionnaire [20]. Stool samples were obtained in the clinic or shipped by patients overnight on wet ice to the Department of Urology, Feinberg School of Medicine. Upon receipt in the lab, written consent forms and questionnaires associated with each individual participant were labeled with the corresponding alpha-numeric identifier, and questionnaire data were entered into a spreadsheet identified only by alpha-numeric identifier. Each stool sample was transferred to a sterile tube containing RNAlater, labeled with the same random alpha-numeric identifier associated with the participant, and stored at –80°. Samples were subsequently shipped on dry ice to the University of Illinois for analyses. There, deep amplicon sequencing was performed in the V3-V5 hypervariable region of the 16S ribosomal RNA gene as previously described, including sequencing on an Illimunia MiSeq V2 sequencer, binning of sequence reads at 97% sequence identity using QIIME and using Galaxy to define abundance of operational taxonomic units (OTUs) [18].

## RESULTS AND DISCUSSION

Fecal stool was obtained from healthy controls and CP/CPPS patients. While age and other demographic data were not significantly different, consistent with their diagnosis CP/CPPS patients’ self-reported symptom scores were significantly higher for all GUPI parameters including overall score, pain, urinary symptoms, and quality of life (Table 1). Globally, principle component analysis (PCA) of 16S rRNA gene sequence analysis revealed overlapping distribution between samples obtained from healthy controls and men diagnosed with CP/CPPS (Fig. 1A). Despite this overlap, CP/CPPS microbiota exhibited altered diversity and richness relative to microbiota of control men (Fig. 1B and C). Overall, diversity scores were shifted higher in CP/CPPS microbiota relative to control microbiota (Fig. 1B). Similarly, overall richness was higher in CP/CPPS microbiota relative controls (Fig. 1C). Whereas OTU richness ranged from 150 – 250 OTUs for control microbiota, a large fraction of CP/CPPS microbiota extended beyond this range, even exceeding 350 OTUs. Together, these findings suggest CP/CPPS microbiota differ from those of healthy controls and are characterized by increased diversity and richness in this cohort.

**Table 1.** CP/CPPS patients and controls

|  | Healthy Controls (n = 9) | CP/CPPS (n = 15) |
| --- | --- | --- |
| Age | **40 ± 15** | **44 ± 14** |
| Sex | **Male** | **Male** |
| Ethnicity | Not Hispanic/Latino: 8/9<br>Hispanic/Latino: 1/9 | Not Hispanic/Latino: 14/15<br>Hispanic/Latino: 1/15 |
| Race | Afr.Am.: 5/9<br>Wht.: 3/9<br>Asian: 1/9 | Afr.Am.: 3/15<br>Wht: 11/15<br>Other: 1/15 |
| Employment Status | Employed: 7/9<br>Unemployed: 2/9 | Employed: 10/15<br>Unemployed: 4/15<br>Retired: 1/15 |
| Family History of PBS/IC | Yes: 0/9 | Yes: 0/15 |
| Family History of CPPS | Yes: 0/9 | Yes: 1/15 |
| Living with Spouse/Partner | Yes: 4/9 | Yes: 3/15 |
| GUPI score total | **0.8 ± 1** | **26 ± 5** |
| GUPI-Pain | **N/A** | **13 ± 2** |
| GUPI-Urinary | **0.4 ± 0.7** | **5 ± 3** |
| GUPI-QOL | **0.3 ± 0.5** | **9 ± 2** |
| Duration of Symptoms (yrs.) | **N/A** | **9.5 ± 8** |
| Genitourinary Disorder | Yes: 0/9 | Yes: 1/15 |
| Respiratory Tract Disorder/Allergies | Yes: 2/9 | Yes: 6/15 |
| Gastrointestinal Disease | Yes: 0/9 | Yes: 2/15 |
| Endocrine/Metabolic Disease | Yes: 0/9 | Yes: 0/15 |
| Hematopoietic/Lymphatic/Infectious Disease | Yes: 0/9 | Yes: 0/15 |
| Psychiatric Disease | Yes: 0/9 | Yes: 5/15 |
| Sexually Transmitted Disease | Yes: 0/9 | Yes: 1/15 |
| Cardiovascular Disease | Yes: 1/9 | Yes: 6/15 |
| Neurologic Disease | Yes: 0/9 | Yes: 2/15 |
| Autoimmune/Other Disorders | Yes: 0/9 | Yes: 1/15 |
| Urological/Gynecologic Surgeries | Yes: 0/9 | Yes: 1/15 |
| Currently Receiving Treatment | Yes: 0/9 | Yes: 4/15 |

**Figure 1.**
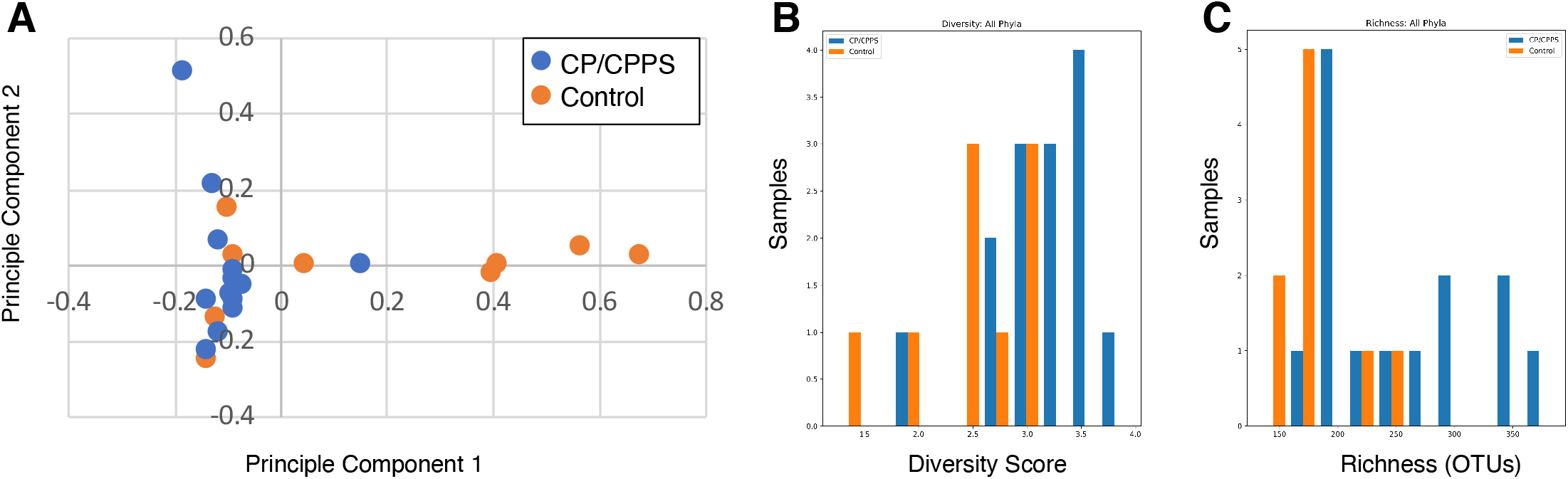
CP/CPPS fecal microbiota differ from healthy male controls. **A**. Principle component analyses of 16S sequence reads from fecal stool samples reveals overlap yet divergence of microbiota of healthy control men from CP/CPPS patients. **B**. CP/CPPS fecal microbiota exhibit increased diversity across all phyla relative to healthy control microbiota. **C**. CP/CPPS fecal microbiota exhibit increased OTU richness relative to healthy control microbiota.

To examine CP/CPPS microbiota more closely, we characterized microbiota richness within phyla. Although several phyla showed no discernable difference, four phyla exhibited a shift toward increased richness in CP/CPPS microbiota (Fig. 2). Increased richness was most evident in the broadest phyla, where CP/CPPS microbiota showed increased richness in Firmicutes and Bacteroidetes (Fig. 2A and B, respectively). In contrast, Proteobacteria and Synergistetes showed increased richness in fewer microbiota (Fig. 2C and D, respectively). Nonetheless, together these data suggest CP/CPPS microbiota are associated with altered richness in both gram-negative and gram-positive phyla.

**Figure 2.**
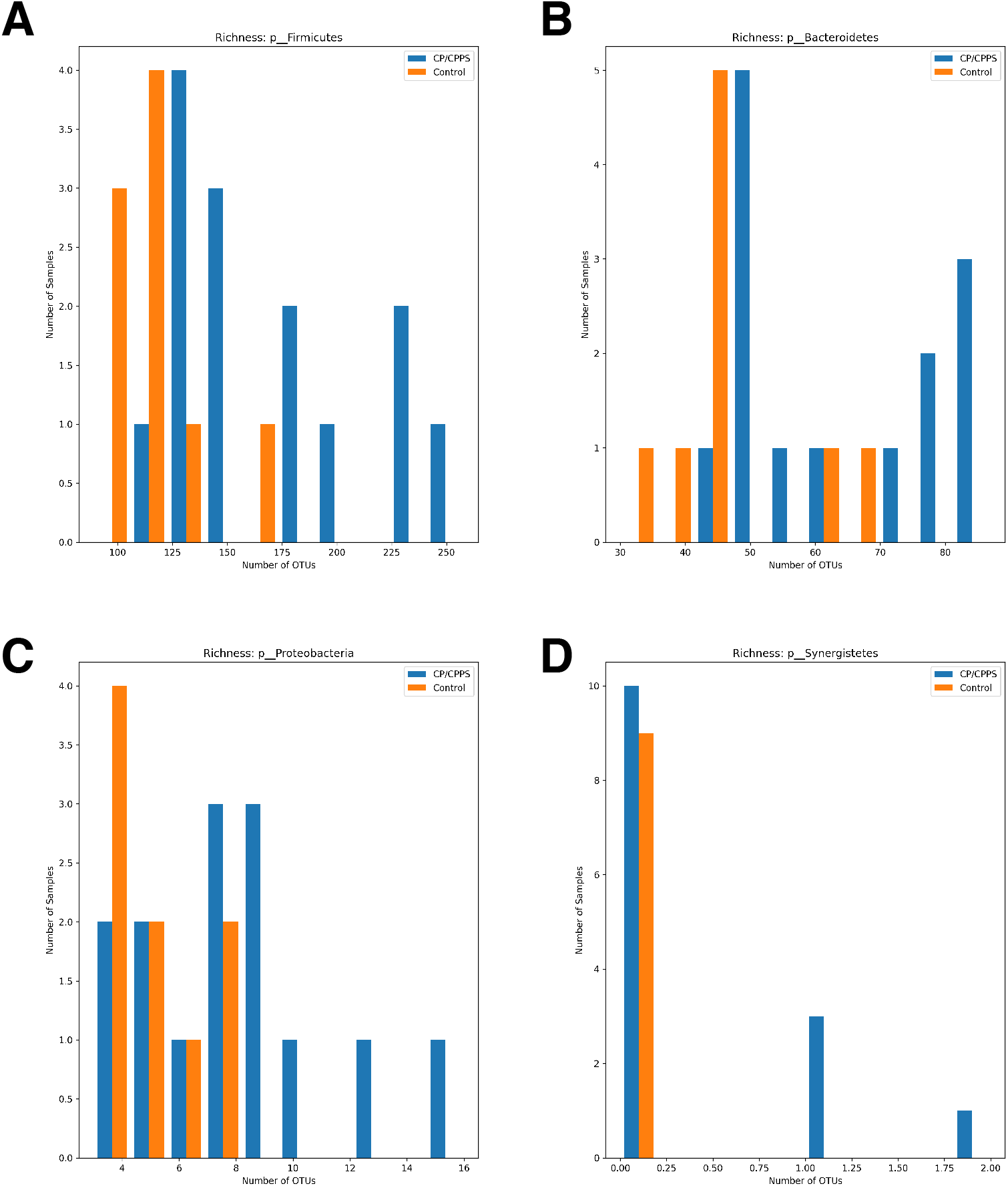
CP/CPPS microbiota are enriched in specific phyla. **A**. CP/CPPS microbiota exhibit increased richness in Firmicutes relative to microbiota of control patients, and **(B)** Bacteriodetes are similarly enriched. Less prevalent phyla of Proteobacteria **(C)** and Synergistetes **(D)** are enriched only in select patients.

Shared among UCPPS is chronic pelvic pain and co-morbid anxiety/depression, despite profound sex differences marked by CP/CPPS occurring exclusively in males and IC/BPS occurring in women in approximately 90% of cases [21]. To identify the potential for shared dysbiosis between CP/CPPS and IC/BPS, we compared microbiota of CP/CPPS with microbiota of IC/BPS patients and for reference included microbiota patients with OAB, LUTS, and MDD (Table 2), diagnoses that share one or more clinical features with UCPPS. PCA plot revealed segregation of CP/CPPS from IC/BPS (Fig. 3). This segregation of CP/CPPS and IC/BPS was not likely due to sex differences because two patients diagnosed with IC/BPS were men (Fig. 3, red oval). Similarly, MDD microbiota were tightly clustered despite including both female and male microbiota. These data suggest that although both CP/CPPS and IC/BPS are associated with fecal dysbiosis, these dysbioses are distinct.

**Table 2.**
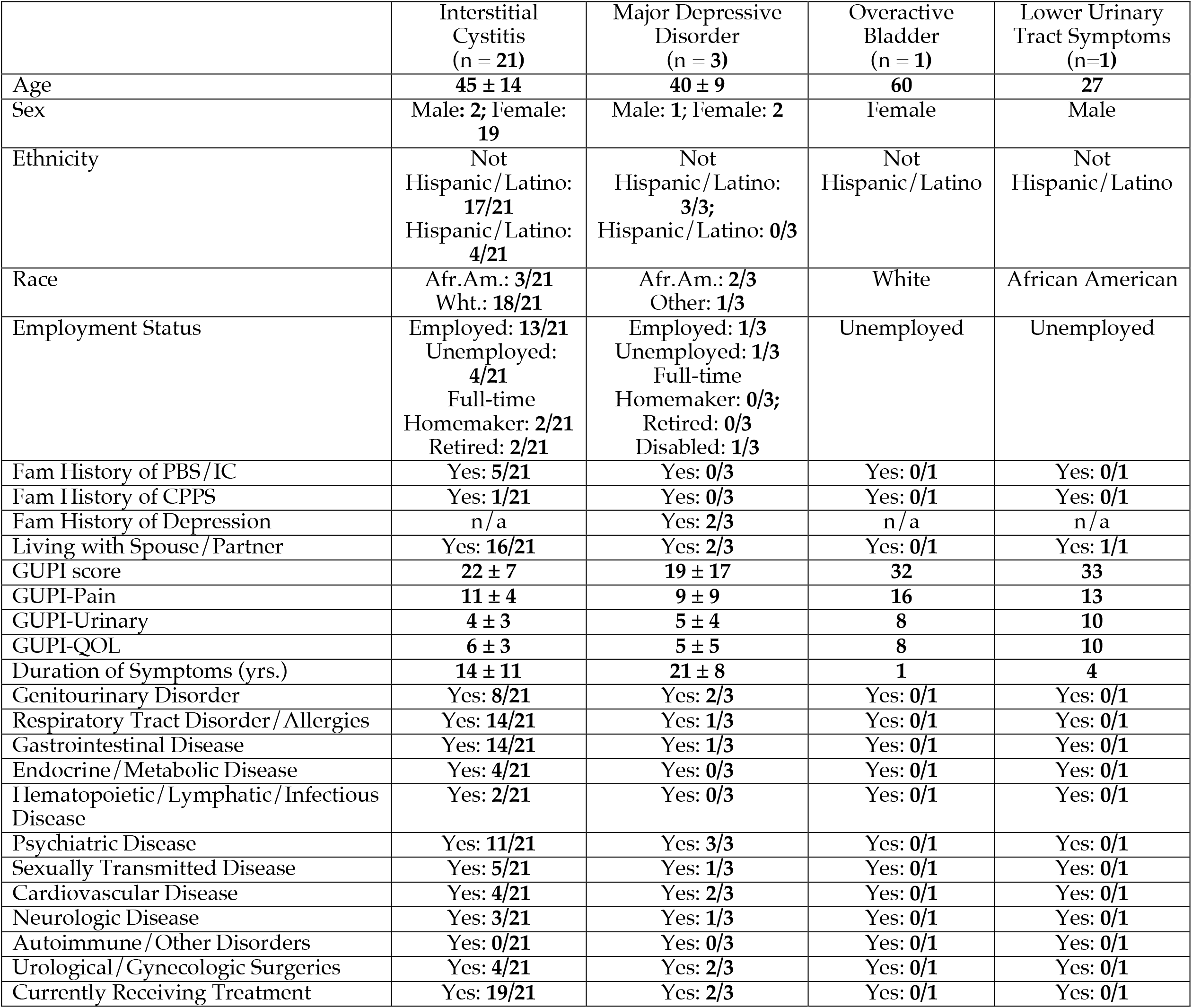
IC, MDD, OAB and LUTS patients

|  | Interstitial Cystitis<br>(n = 21) | Major Depressive Disorder<br>(n = 3) | Overactive Bladder<br>(n = 1) | Lower Urinary Tract Symptoms<br>(n=1) |
| --- | --- | --- | --- | --- |
| Age | 45 ± 14 | 40 ± 9 | 60 | 27 |
| Sex | Male: 2; Female: 19 | Male: 1; Female: 2 | Female | Male |
| Ethnicity | Not Hispanic/Latino: 17/21<br>Hispanic/Latino: 4/21 | Not Hispanic/Latino: 3/3;<br>Hispanic/Latino: 0/3 | Not Hispanic/Latino | Not Hispanic/Latino |
| Race | Afr.Am.: 3/21<br>Wht.: 18/21 | Afr.Am.: 2/3<br>Other: 1/3 | White | African American |
| Employment Status | Employed: 13/21<br>Unemployed: 4/21<br>Full-time Homemaker: 2/21<br>Retired: 2/21 | Employed: 1/3<br>Unemployed: 1/3<br>Full-time Homemaker: 0/3;<br>Retired: 0/3<br>Disabled: 1/3 | Unemployed | Unemployed |
| Fam History of PBS/IC | Yes: 5/21 | Yes: 0/3 | Yes: 0/1 | Yes: 0/1 |
| Fam History of CPPS | Yes: 1/21 | Yes: 0/3 | Yes: 0/1 | Yes: 0/1 |
| Fam History of Depression | n/a | Yes: 2/3 | n/a | n/a |
| Living with Spouse/Partner | Yes: 16/21 | Yes: 2/3 | Yes: 0/1 | Yes: 1/1 |
| GUPI score | 22 ± 7 | 19 ± 17 | 32 | 33 |
| GUPI-Pain | 11 ± 4 | 9 ± 9 | 16 | 13 |
| GUPI-Urinary | 4 ± 3 | 5 ± 4 | 8 | 10 |
| GUPI-QOL | 6 ± 3 | 5 ± 5 | 8 | 10 |
| Duration of Symptoms (yrs.) | 14 ± 11 | 21 ± 8 | 1 | 4 |
| Genitourinary Disorder | Yes: 8/21 | Yes: 2/3 | Yes: 0/1 | Yes: 0/1 |
| Respiratory Tract Disorder/Allergies | Yes: 14/21 | Yes: 1/3 | Yes: 0/1 | Yes: 0/1 |
| Gastrointestinal Disease | Yes: 14/21 | Yes: 1/3 | Yes: 0/1 | Yes: 0/1 |
| Endocrine/Metabolic Disease | Yes: 4/21 | Yes: 0/3 | Yes: 0/1 | Yes: 0/1 |
| Hematopoietic/Lymphatic/Infectious Disease | Yes: 2/21 | Yes: 0/3 | Yes: 0/1 | Yes: 0/1 |
| Psychiatric Disease | Yes: 11/21 | Yes: 3/3 | Yes: 0/1 | Yes: 0/1 |
| Sexually Transmitted Disease | Yes: 5/21 | Yes: 1/3 | Yes: 0/1 | Yes: 0/1 |
| Cardiovascular Disease | Yes: 4/21 | Yes: 2/3 | Yes: 0/1 | Yes: 0/1 |
| Neurologic Disease | Yes: 3/21 | Yes: 1/3 | Yes: 0/1 | Yes: 0/1 |
| Autoimmune/Other Disorders | Yes: 0/21 | Yes: 0/3 | Yes: 0/1 | Yes: 0/1 |
| Urological/Gynecologic Surgeries | Yes: 4/21 | Yes: 2/3 | Yes: 0/1 | Yes: 0/1 |
| Currently Receiving Treatment | Yes: 19/21 | Yes: 2/3 | Yes: 0/1 | Yes: 0/1 |

**Figure 3.**
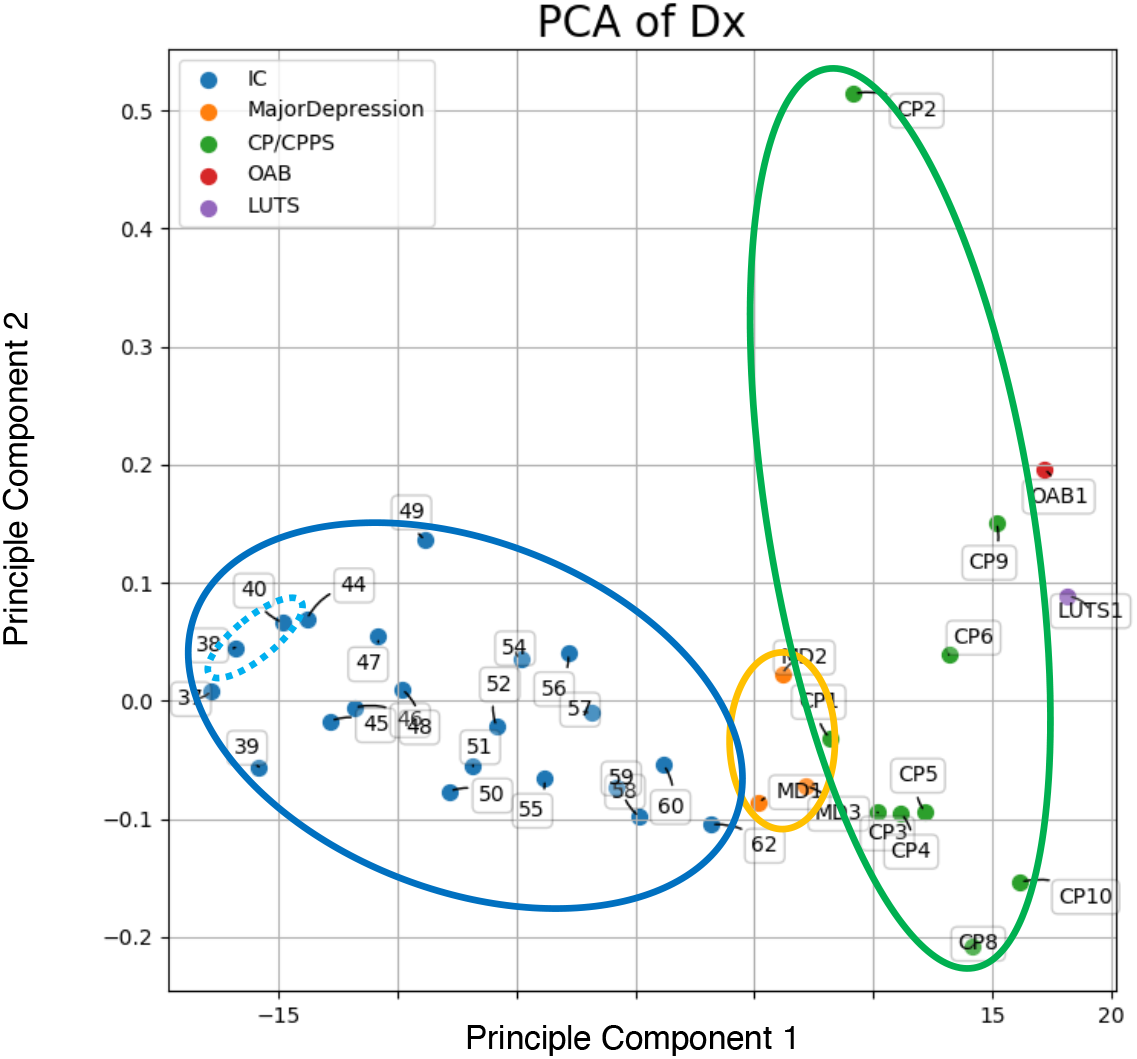
CP/CPPS and IC/BPS microbiota are distinct. PCA analysis reveals segregation of clustered CP/CPPS microbiota (green oval) from clustered IC/BPS microbiota (blue oval). Two male IC/BPS microbiota are indicated (dashed blue oval). Male and female microbiota of patients with depression cluster (orange oval).

Here we characterized fecal microbiota of CP/CPPS patients. Like prior studies, we identified altered microbiota consistent with dysbiosis in men with CP/CPPS. CP/CPPS microbiota in our cohort were associated with overall increased diversity and increased richness (Fig. 1). However, this is in contrast to prior studies that observed dysbiosis marked by reduced microbiota diversity among CP/CPPS patients where samples were rectal microbiota harvested on a glove tip [19]. Although we examined participant-collected fecal stool, recent work found that microbiota harvested via rectal swab, glove tip, and participant-collected stool yielded similar results [22], suggesting sampling method does not underlie the differences between our findings and those of Shoskes and colleagues. Rather it is more likely that differences between this study and that of Shoskes reflect differences in study cohorts including but not limited to sample size, age, and geographic effects on diet. Nonetheless, the fact that both studies identify dysbiosis among CP/CPPS patients yet with distinct differences suggests the need to expanded studies of the UCPPS microbiome.

We previously identified dysbiosis in IC/BPS patients as well as associated metabolomic changes in fecal stool [18]. In this pilot study of UCPPS microbiota, we compared IC/BPS with CP/CPPS (Fig. 3). Surprisingly, IC/BPS and CP/CPPS microbiota diverged. Other studies have yet to find strong sex effects on human microbiota where sex has been found to explain only 0.5% of variation in gut microbiota, less than for other factors including diet and medication (reviewed in [23]). Moreover, in this study two male IC/BPS microbiota clustered among female IC/BPS microbiota and separate from CP/CPPS, while male and female microbiota of depression patients were tightly grouped. Together, our findings and review of sex effects on microbiota suggest that the differences between CP/CPPS and IC/BPS microbiota are due to factors other than sex. Instead, we speculate differences in CP/CPPS and IC/BPS microbiota reflect differences in UCPPS etiology or pathogenic mechanism between CP/CPPS and IC/BPS, despite overlapping symptom complexes.

In conclusion, consistent with prior studies, we find CP/CPPS patients exhibit gut dysbiosis. However, CP/CPPS microbiota are dissimilar from IC/BPS microbiota not attributed to sex, suggesting distinct factors contributing to dysbiosis among UCPPS patients.

## Data Availability

All raw and edited data is available upon request from Dr. Klumpp at Northwestern University. This data is not available online.

## ACKNOWLEDGEMENTS

We thank Mr. Lucius Robinson for exemplary work recruiting and screening patients.

